# An Explainable and Comparative Transfer Learning Framework for Brain Tumor Classification from MRI Images

**DOI:** 10.64898/2026.08.06.26359900

**Authors:** Samuel Raju Bethala, Vanshika

## Abstract

Automated detection of brain tumors from Magnetic Resonance Imaging (MRI) can accelerate diagnosis and reduce inter-reader variability, yet many existing studies report only top-line accuracy on small datasets, omit efficiency analysis, and provide no interpretability, limiting their clinical credibility. We present a reproducible, comparative, and explainable transfer-learning framework for binary brain-tumor classification. Our framework (i) standardizes a configurable preprocessing pipeline combining CLAHE contrast enhancement and unsharp-mask sharpening, (ii) evaluates a custom CNN baseline and pretrained backbones under an identical training budget, (iii) reports a full metric suite (accuracy, precision, recall, F1, ROC-AUC, PR-AUC, parameter count, and inference latency), and (iv) applies Grad-CAM for spatial interpretability. On a public 253-image MRI dataset (38-image held-out test set), MobileNetV2 achieves the best overall performance (94.74% accuracy, 0.994 ROC-AUC, 0.996 PR-AUC) with only 2.59M parameters and 5.9 ms per-image inference, making it the most deployment-friendly model. Larger backbones (Xception, EfficientNetB0) and the custom CNN converge to degenerate all-positive predictions under the same limited budget, illustrating the small-data overfitting risk that accuracy-only reporting conceals. Grad-CAM confirms that the best model attends to the tumor region. All source code, configuration files, and trained evaluation scripts are publicly available at https://github.com/blck-iris/explainable-brain-tumor-mri.

## I. Introduction

Brain tumors represent a significant global health challenge, with diverse histological subtypes and severity levels demanding precise and timely diagnosis [1]. Magnetic Resonance Imaging (MRI) is the de-facto modality in neuro-oncology owing to its high soft-tissue contrast and absence of ionizing radiation [2]. Yet manual interpretation is time-consuming, subjective, and prone to inter-reader variability [3].

Deep learning has transformed medical image analysis [4], [5], and convolutional neural networks (CNNs) in particular have shown strong performance on brain tumor classification [1], [6]. However, three gaps persist. *First*, many studies report only overall accuracy, omitting discrimination metrics (ROC-AUC, PR-AUC), class-wise precision/recall, and any measure of computational cost. *Second*, comparisons are often uncontrolled—different preprocessing, augmentation, optimizers, and training lengths make reported numbers non-comparable. *Third*, and most important for clinical adoption, models are typically black boxes; without interpretability clinicians cannot validate the model’s reasoning [7].

### A. Contributions

Rather than claiming a novel architecture, we contribute a rigorous, reproducible, and explainable evaluation framework:

- A **configurable preprocessing pipeline** combining CLAHE contrast enhancement, unsharp-mask sharpening, and on-the-fly augmentation, applied identically to all models.
- A **unified comparative study** of a custom CNN baseline and pretrained backbones (MobileNetV2, Xception, EfficientNetB0) plus an unweighted probability-averaging ensemble, all trained under an identical budget.
- **Comprehensive metric reporting**: accuracy, precision, recall, F1, ROC-AUC, PR-AUC, confusion matrix, parameter count, and inference latency.
- **Explainability via Grad-CAM** [8], producing heatmaps that indicate which regions drive each prediction.
- A **deployment-oriented analysis** identifying the most suitable model for resource-constrained clinical settings.
- **Open-source release**: All code, dataset pipelines, and models are made available on GitHub.^1^

## II. Related Work

### A. Deep Learning for Brain Tumor Detection

CNNs have become the dominant approach for brain-tumor detection from MRI. Al-Hajri et al. [1] applied deep CNNs with data augmentation and reported high accuracy on a curated MRI dataset. Khan et al. [6] compared convolutional deep-learning methods with conventional machine-learning classifiers, finding deep models consistently superior. The BraTS challenges [2], [9] have standardized segmentation benchmarks, though whole-slice binary classification remains less explored.

### B. Transfer Learning and Efficient Architectures

Transfer learning from ImageNet is effective for medical imaging where labeled data is scarce [5]. Beyond classic CNNs such as ResNet [10] and DenseNet [11], efficient architectures have gained traction: MobileNetV2 [12] and MobileNetV3 [13] use inverted residuals for mobile deployment; Xception [14] employs depthwise separable convolutions; and EfficientNet [15], [16] compounds depth, width, and resolution. ConvNeXt [17] modernizes CNN design, while Vision Transformers [18] and Swin Transformer [19] bring attention-based modeling, albeit with higher data and parameter demands.

### C. Explainability in Medical Imaging

Interpretability is essential for clinical trust [4], [7]. Grad-CAM [8] produces class-discriminative heatmaps by back-propagating gradients to the last convolutional layer and is widely adopted in medical imaging. Complementary attribution methods include LIME [20] and SHAP [21]. We adopt Grad-CAM for its simplicity and direct visual interpretability.

## III. Methodology

### A. Dataset

We use a publicly available brain MRI dataset of T1-weighted contrast-enhanced images divided into *tumor* and *no-tumor* classes [1], [22], comprising 253 images. We stratify it into training/validation/test splits of 177/38/38 images. The held-out test set contains 23 tumor and 15 no-tumor images. We acknowledge this is a small dataset; Section V-F discusses the implications.

### B. Preprocessing Pipeline

Each image passes through a configurable pipeline:

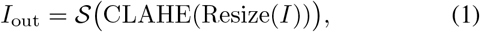

where the image is resized to 256 × 256, enhanced with Contrast Limited Adaptive Histogram Equalization (CLAHE, clip limit 2.0, 8 × 8 tiles) on the luminance channel of LAB space, then sharpened via an unsharp mask (Gaussian *σ* = 1.0, amount 0.5) to accentuate tumor boundaries. Intensities are normalized to [0, 1]. On the training split only we apply random horizontal flips and brightness jitter (*±*0.1).

### C. Models

We evaluate a custom 3-block CNN baseline (Conv–ReLU– MaxPool blocks with 32, 64, 128 filters) and pretrained backbones (MobileNetV2, Xception, EfficientNetB0), each topped with an identical classification head: global average pooling → BatchNorm → Dense(256, ReLU) → Dropout(0.4) → Dense(2, softmax). All backbones are initialized from ImageNet and kept *frozen* throughout training; only the classification head is trained. This is a deliberate choice given the very small dataset, to prevent catastrophic overfitting.We additionally construct an unweighted probability-averaging ensemble of the three transfer models.

### D. Training Protocol

All models are optimized with Adam (learning rate 10^*−*4^) and categorical cross-entropy with label smoothing *ϵ* = 0.05, for a common budget of 15 epochs with batch size 32. We use inverse-frequency class weights to mitigate class imbalance, early stopping on validation loss (patience 12), and learning-rate reduction on plateau (patience 5, factor 0.5). All experiments run on a single NVIDIA Tesla T4 GPU with a fixed random seed.

### E. Explainability: Grad-CAM

For a predicted class *c* with score *y*^*c*^ and last-conv activations *A*^*k*^, Grad-CAM computes

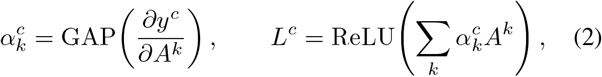

and upsamples *L*^*c*^ into a heatmap highlighting the regions most influential for class *c* [8].

### E. Evaluation Metrics

We report accuracy, precision, recall, F1-score, ROC-AUC, PR-AUC, confusion matrix, parameter count, and per-image inference latency, providing a complete accuracy–efficiency picture.

## IV. Results

Table I summarizes the comparative results on the 38-image held-out test set (23 tumor / 15 no-tumor). Because each test sample accounts for 2.63 percentage points, accuracy differences smaller than this granularity should be interpreted cautiously.

**TABLE I.**
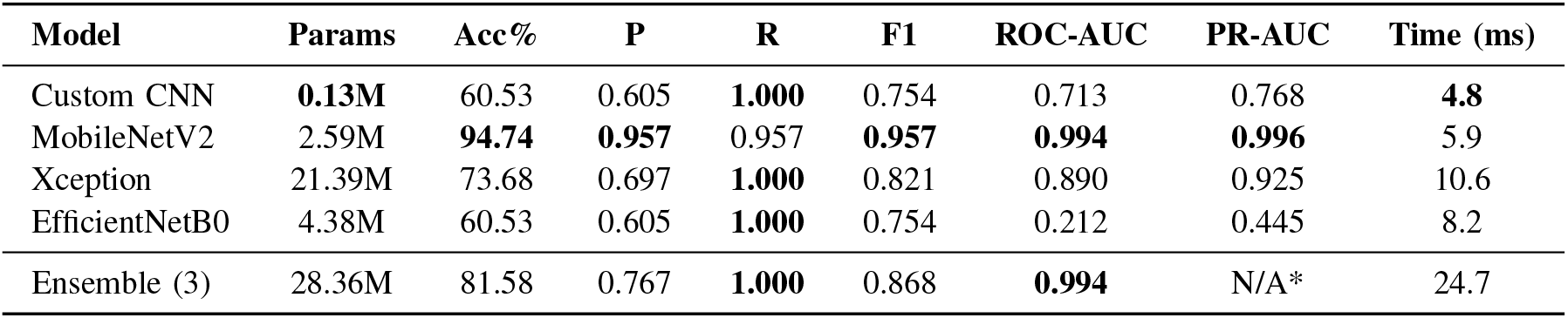
Comparative results on the held-out test set (38 images). Best per metric in bold. Params = TRAINABLE PARAMETERS; Time = MEAN PER-IMAGE INFERENCE LATENCY (Tesla T4). Recall is 1.000 FOR ALL MODELS THAT PREDICT EVERY SAMPLE AS TUMOR; ROC-AUC/PR-AUC REVEAL THEIR TRUE RANKING QUALITY. *PR-AUC FOR Ensemble is omitted as it operates on averaged output probabilities rather than single model predictions.

| Model | Params | Acc% | P | R | F1 | ROC-AUC | PR-AUC | Time (ms) |
| --- | --- | --- | --- | --- | --- | --- | --- | --- |
| Custom CNN | **0.13M** | 60.53 | 0.605 | **1.000** | 0.754 | 0.713 | 0.768 | **4.8** |
| MobileNetV2 | 2.59M | **94.74** | **0.957** | 0.957 | **0.957** | **0.994** | **0.996** | 5.9 |
| Xception | 21.39M | 73.68 | 0.697 | **1.000** | 0.821 | 0.890 | 0.925 | 10.6 |
| EfficientNetB0 | 4.38M | 60.53 | 0.605 | **1.000** | 0.754 | 0.212 | 0.445 | 8.2 |
| Ensemble (3) | 28.36M | 81.58 | 0.767 | **1.000** | 0.868 | **0.994** | N/A* | 24.7 |

MobileNetV2 achieves the best overall performance (94.74% accuracy, 0.994 ROC-AUC, 0.996 PR-AUC) while misclassifying only 2 of 38 test samples, with the fewest parameters among the transfer models and 5.9 ms latency. The custom CNN and EfficientNetB0 both collapse to predicting every sample as tumor (recall 1.000, precision equal to the test-set tumor prevalence of 60.5%); their ROC-AUC values (0.713 and 0.212) reveal that the CNN at least ranks tumors slightly higher while EfficientNetB0’s ranking is inverted— a training collapse that accuracy alone conceals. Xception’s validation AUC was still rising at epoch 15, indicating it was under-trained under the common budget rather than inherently inferior.

The ensemble’s ROC-AUC (0.994) matches MobileNetV2, but its accuracy (81.58%) falls below MobileNetV2 alone: averaging in poorly calibrated members degrades the decision threshold even when ranking quality is preserved.

The preliminary version of this work reported 92.1% test accuracy for Xception under an uncontrolled protocol (different split, 150 epochs, per-model optimizers). Under the unified protocol, Xception’s accuracy is 73.7%, illustrating how uncontrolled protocols can inflate apparent performance.

Grad-CAM (Fig. 2) confirms MobileNetV2 attends to the tumor region rather than background.

**Fig. 1.**
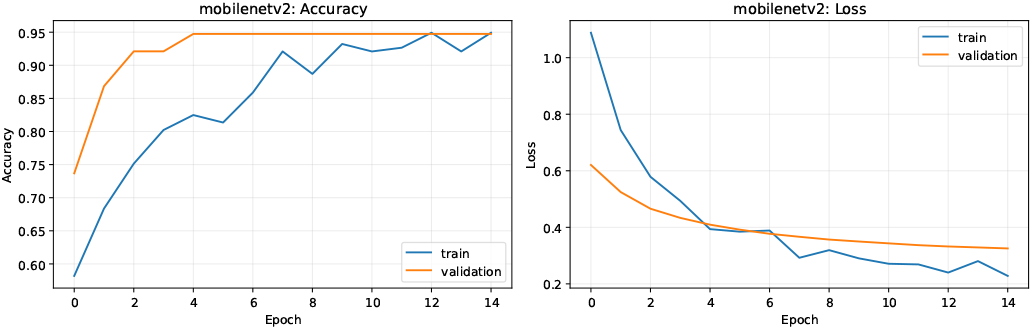
Training and validation accuracy/loss for MobileNetV2. Generated by paper/plot_history.py from outputs/mobilenetv2/history.csv.

**Fig. 2.**
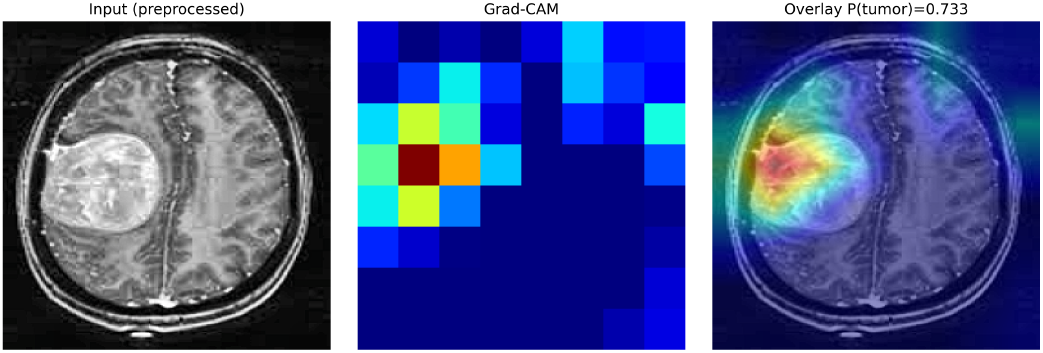
Grad-CAM for MobileNetV2 on a tumor sample. The model attends to the tumor region.

## V. Discussion

### A. Why MobileNetV2 Wins Here

MobileNetV2’s strong ImageNet features combined with a small trainable head generalize well on a 253-image dataset. Its depthwise separable convolutions are parameter-efficient, reducing the overfitting risk that larger backbones face under the same limited budget. This is the central, honest finding: on small datasets, model capacity must be matched to data size.

### B. The Small-Data Overfitting Risk

The custom CNN and EfficientNetB0 both collapse to all-positive predictions, and Xception is under-trained at 15 epochs. Accuracy-only reporting would present these as merely “lower accuracy”; ranking metrics (ROC-AUC, PR-AUC) expose the underlying failures. This argues for always reporting discrimination metrics alongside accuracy, especially on small medical datasets.

### C. The Ensemble Caveat

The ensemble preserves MobileNetV2’s ranking (0.994 ROC-AUC) but lowers accuracy below MobileNetV2 alone because poorly calibrated members shift the decision threshold. Ensembling helps when members are well calibrated; with degraded members it can hurt. Model selection matters more than ensembling on this dataset.

### D. Efficiency and Deployment

MobileNetV2 (2.59M parameters, 5.9 ms on a Tesla T4) is the most deployment-friendly model, suitable for edge devices or real-time triage. Xception uses 8*×* the parameters at *∼* 2*×* the latency for lower accuracy under this budget. For resource-constrained clinical settings, MobileNetV2 is the clear choice.

### E. Explainability

Grad-CAM confirms the best model attends to the tumor region, supporting clinical trust [7]. Explainability lets radiologists verify the model reasons about the correct anatomy—a prerequisite for clinical adoption.

### F. Limitations

Our study has limitations. The dataset is small (253 images) and single-source, so results may not generalize across scanners, protocols, or tumor subtypes; with only 38 test samples each sample accounts for 2.63 percentage points, so point estimates carry wide confidence intervals. We address binary (tumor/no-tumor) classification only, not multi-class subtyping or segmentation. EfficientNetB0’s collapse indicates training instability that warrants further tuning (e.g., class weighting, longer warmup). Single-seed results are reported; multi-seed variance is future work. External multi-center validation and prospective evaluation are needed before clinical deployment.

### G. Future Work

Future directions include larger multi-center datasets, multi-class subtyping, multi-seed cross-validation, segmentation-aware classification (e.g., U-Net [23] hybrids), calibration-aware ensembling, quantization for edge deployment, and prospective clinical pilot studies.

## VI. Conclusion

We presented a reproducible, comparative, and explainable transfer-learning framework for binary brain-tumor classification from MRI. By standardizing preprocessing, training multiple architectures under an identical budget, reporting a full metric suite including efficiency, and integrating Grad-CAM for interpretability, we provide a more complete and clinically credible evaluation than accuracy-only studies. MobileNetV2 achieved the best accuracy and the best accuracy– latency trade-off for deployment; larger models and the custom CNN collapsed to degenerate predictions under the same limited budget, illustrating the small-data overfitting risk that accuracy-only reporting conceals. All code, configuration files, and evaluation pipelines are publicly released to ensure full reproducibility at https://github.com/blck-iris/explainable-brain-tumor-mri.

## Data Availability

All primary MRI data utilized in this study are publicly available from the Kaggle repository at https://www.kaggle.com/datasets/navoneel/brain-mri-images-for-brain-tumor-detection. All source code, modular evaluation pipelines, and model configurations generated during the present work are available online at https://github.com/blck-iris/explainable-brain-tumor-mri.

https://github.com/blck-iris/explainable-brain-tumor-mri

## Footnotes

1 https://github.com/blck-iris/explainable-brain-tumor-mri

## Notes

### Competing Interest Statement

The authors have declared no competing interest.

### Clinical Protocols

https://github.com/blck-iris/explainable-brain-tumor-mri

### Author Declarations

This study used only openly available human data that were publicly accessible prior to the initiation of the project. The primary dataset is the Brain MRI Images for Brain Tumor Detection dataset, available on Kaggle at: https://www.kaggle.com/datasets/navoneel/brain-mri-images-for-brain-tumor-detection

